# Financial Incentives and Healthcare Provision: Evidence from an Experimental *Aedes aegypti* Control Programme in Brazil

**DOI:** 10.1101/2021.03.10.21252321

**Authors:** Danilo Freire, Umberto Mignozzetti

## Abstract

**Background:** Mosquito control is the most effective means of reducing *Aedes aegypti* infections worldwide. In many developing countries, however, vector management programmes fail to reach their goals due to low worker productivity. Research suggests that financial incentives may increase the productivity of health personnel, yet there is little evidence about the impact of monetary rewards on *A. aegypti*-reduction strategies. We evaluated whether individual and collective financial incentives improve the performance of healthcare workers fighting *A. aegypti*, as well as their effect on city-level numbers of dengue hospitalisations.

**Methodology/Principal findings:** We hired and trained subjects to visit households, find *A. aegypti* breeding sites, and eliminate mosquito larvae in the city of Rio Verde, Brazil. We randomly assigned workers into three groups. The control group received a flat compensation for their tasks, while workers in the two treatment groups received individual and collective monetary bonuses, respectively. Financial rewards increased the number of cleaned breeding sites in both treatment groups (individual and team bonuses), and the collective treatment also improved larvae extermination. The intervention lowered dengue hospitalisations in 10.3%, but the result was not consistent across all model specifications.

**Conclusions/Significance:** *A. aegypti* control programmes may benefit from alternative compensation schemes, especially when provided to teams. For this strategy to succeed, financial incentives have to be distributed widely as their aggregate effect is limited. More research is needed to assess whether higher worker productivity decreases dengue hospitalisations.

**Author Summary:** Diseases transmitted by the *Aedes aegypti* mosquito, such as chikungunya, dengue, yellow fever, and Zika, continue to affect thousands of people per year. As there are no safe vaccines for most of these infections, insecticide spraying and breeding site elimination are the best means to fight the mosquito. In several developing countries, which host the majority of *A. aegypti* infections, anti-mosquito campaigns are carried out inconsistently, thus it is crucial to find ways to improve the productivity of healthcare workers in charge of these tasks. We designed a randomised field experiment that provided individual and collective financial incentives to healthcare agents in a Brazilian city, and we tested the effect of monetary rewards on their productivity and on city-level dengue hospitalisations. We find that financial bonuses improved the number of cleaned breeding sites in both treatment groups (individual and team incentives) and that the collective treatment also improved larvae extermination. The impact of our treatment on city-level hospitalisations was not consistent across all specifications. In sum, financial incentives may be used to boost field productivity in anti-*A. aegypti* programmes, but further research is required to evaluate how healthcare worker productivity impacts dengue outcomes.

## 1 Introduction

The *Aedes aegypti* mosquito is the major worldwide vector of several arboviral diseases, including chikungunya, dengue, yellow fever, and Zika [1, 2, 3, 4, 5]. Over the last decades, arboviral incidence has increased exponentially, with estimates suggesting that dengue is endemic in over 100 countries and causes 400 million infections and 22 thousand deaths each year [6, 7]. Since there are no safe vaccines for most *Aedes*-transmitted diseases [8], we have seen a number of dengue and Zika outbreaks in Asia, the Caribbean, South America, and the United States [9, 4]. Chikungunya virus, previously endemic only to Africa, has already been reported in every region of the world [10, 11, 12].

Mosquito control is the primary strategy to prevent *Aedes*-borne infections [13, 14]. The World Health Organization recommends the cleaning of containers and the use of insecticide to reduce vector breeding, especially in settings where human-vector contacts are frequent, such as schools, hospitals, and workplaces [15, 16]. However, breeding site elimination and insecticide spraying are labour-intensive processes, which are often carried out inconsistently by developing states [17, 18, 15]. In this regard, improving the performance of their healthcare agencies is essential to the effectiveness of anti-*Aedes* programmes.

Research shows that financial incentives may increase the productivity of healthcare workers [19, 20, 21]. Nevertheless, systematic reviews indicate that the internal validity of the available evidence is limited due to methodological shortcomings, as several studies suffer from selection bias and inadequate experimental blinding [22, 23, 24]. The reviews also point out that it remains unclear which rewarding schemes have a stronger impact on healthcare worker performance [25] and whether monetary incentives have any sustained effects on patient outcomes [26, 27].

In this paper, we explore how financial incentives affect the productivity of healthcare personnel involved in *A. aegypti* control and how they impact dengue hospitalisations. We ran a randomised field experiment in the city of Rio Verde, Brazil, which has experienced a peak in dengue infections in recent years. We hired and trained teams of healthcare workers to perform simplified procedures designed to exterminate *A. aegypti* breeding sites. Our experiment consisted of two treatment arms plus a control condition. We offered individual or collective bonuses to those allocated in the two treatment groups, while the control group received a fixed compensation for completing their tasks. We then measured how the different reward schemes affected the number of breeding sites discovered, larvae exterminated, and city-wide dengue incidence.

We find that monetary rewards increase the number of cleaned breeding sites in both treatment conditions (individual and team bonuses), and that collective financial incentives also improve larvae extermination. Our results show that the control group has a high number of houses visited in less than two minutes apart from each other, which suggests that workers may cheat in the absence of monetary incentives. When we combine both treatments, we also find that the intervention decreases dengue incidence in 10.3%, but the results are not robust. In summary, we provide experimental evidence that performance bonuses enhance public service delivery and that small financial incentives may improve patient outcomes and reduce hospitalisations in *Aedes*-endemic countries.

## 2 Methods

We conducted a randomised field trial in Rio Verde, a municipality in the Brazilian Midwestern state of Goiás. In 2017, Rio Verde had 229,000 inhabitants and 1,156 confirmed dengue cases, one of highest counts in the state [28]. We used Facebook advertisements to recruit potential participants. The advertisements directed applicants to Google Forms, where we asked for their contact information. We sent a pre-treatment demographic survey to every applicant who provided a valid email address. At this stage, we excluded all individuals under 18 years old and invited the remaining applicants to our training session, which we ran on the 4^th^ of May, 2018. The meeting involved a lecture on *A. aegypti* vector biology and practical instructions on how to find and exterminate mosquito breeding sites. Those who did not attend the session were also excluded from the study.

Table 1 displays the attrition rates and the covariance balance tests for our experiment. The last two tables report the values for our territorial assignment before and after applying propensity score matching [29] on census tract characteristics. We observe no significant differences regarding attrition rates, indicating that the treatment was correctly randomised. Participant demographics are also similar across all groups, which suggests that the matching technique successfully balanced the territorial assignment.

**Table 1:** Covariate Balance on Individual Characteristics

| Variable | Attrition | Control | Individual | Collective | F-Statistic | P-Value |
| --- | --- | --- | --- | --- | --- | --- |
| Panel A: Attrition Pattern Test |  |  |  |  |  |  |
| Age | 26.62 | 24.74 | 26.02 | 26.19 | 1.056 | 0.367 |
| Female | 0.73 | 0.81 | 0.72 | 0.7 | 0.723 | 0.539 |
| Has Vehicle | 0.37 | 0.29 | 0.34 | 0.41 | 0.666 | 0.574 |
| N | 263 | 58 | 64 | 74 |  |  |
| Variable |  | Control | Individual | Collective | F-Statistic | P-Value |
| Panel B: Covariate Balance on Individual Characteristics |  |  |  |  |  |  |
| Age |  | 24.74 | 26.02 | 26.19 | 0.751 | 0.473 |
| Altruist |  | 0.62 | 0.66 | 0.61 | 0.176 | 0.839 |
| Religiosity |  | 0.52 | 0.38 | 0.43 | 1.257 | 0.287 |
| Pol. Engagement |  | 0.64 | 0.66 | 0.7 | 0.335 | 0.716 |
| Soc. Engagement |  | 0.55 | 0.59 | 0.64 | 0.467 | 0.627 |
| Above Two Min. Wage |  | 0.34 | 0.38 | 0.28 | 0.671 | 0.513 |
| Facebook Popularity |  | 0.53 | 0.58 | 0.49 | 0.576 | 0.563 |
| Panel C: Unmatched Covariate Balance on Territorial Assignment |  |  |  |  |  |  |
| Number of Houses |  | 276.41 | 243.58 | 255.2 | 2.368 | 0.098 |
| Number of Households |  | 885.67 | 750.34 | 785.18 | 3.765 | 0.026 |
| Avg. Household Size |  | 3.21 | 3.07 | 3.07 | 6.566 | 0.002 |
| Log Avg. Income |  | 6.47 | 6.86 | 6.85 | 9.213 | 0 |
| Cases Before Treatment |  | 13.78 | 5.94 | 4.66 | 15.638 | 0 |
| Panel D: Matched Covariate Balance on Territorial Assignment |  |  |  |  |  |  |
| Number of Houses |  | 276.41 | 282.04 | 283.5 | 0.178 | 0.837 |
| Number of Households |  | 885.67 | 903.04 | 907.14 | 0.154 | 0.858 |
| Avg. Household Size |  | 3.21 | 3.21 | 3.2 | 0.019 | 0.981 |
| Log Avg. Income |  | 6.47 | 6.45 | 6.46 | 0.071 | 0.931 |
| Cases Before Treatment |  | 13.78 | 10.24 | 10.91 | 1.791 | 0.171 |
| Pairs |  | 29 | 32 | 37 |  |  |
| N |  | 58 | 64 | 74 |  |  |

The intervention took place on the 5^th^ of May, 2018. We randomly assigned subjects into three groups and directed each group to separate headquarters to avoid spillovers. In the control group (n = 58), we told participants that they would receive a flat payment regardless of their performance (BRL 110 / USD 25 per subject). In the individual treatment group (n = 64), we informed participants that we would rank each one of them separately and double the compensation for those who performed above the sample median (BRL 220 / USD 50 per subject). In the collective treatment group (n = 74), we explained that we would rank the performance of each group and double the payment for those teams whose results were higher than the sample median (BRL 220 / USD 50 per subject). Participants worked in pairs and were aware of other groups, but they did not know about the different treatment conditions. The teams received a leaflet with a map of the area they should cover. Each route included around 120 houses in two or three blocks.

A typical household visit consisted of the following steps. First, participants rang the doorbell and explained that the municipality had experienced a dengue fever outbreak. They instructed dwellers on how to lower dengue incidence and handed them a leaflet with information about dengue prevention practices. Then, subjects asked whether they were allowed to inspect the resident’s house yard. If granted permission, they entered the household and searched for clean breeding sites, such as pots filled with clear water, and for recipients containing larvae. Upon exterminating a breeding site (pouring out the accumulated water, for example), subjects had to report their task by taking a photo on their cellphones. When they discovered larvae, participants had to record a short video showing the larvae before exterminating them. At the end of each visit, subjects had to take a picture of the household to account for their presence in the location.

This study evaluates two outcomes. The first outcome is the field productivity of healthcare providers. We measure worker productivity by coding cellphone data collected during the intervention into four indicators: 1) the number of breeding sites removed and cleaned; 2) the number of exterminated larvae; 3) the number of houses visited; 4) the number of houses visited within less than two minutes. We analyse the data using ordinary least squares with robust standard errors [30]. We estimate the statistical power of the study assuming a treatment effect of 0.15 standard deviations.

The second outcome of interest is dengue incidence in the municipality of Rio Verde measured 4, 8, 12, and 16 days after the intervention. The Rio Verde Mayor’s Office provided us with its annual healthcare report, which contains the dates and personal information of patients who had dengue fever and received hospital treatment from January to September 2018. We georeferenced the home addresses of the patients and compared the incidence map with the distribution of our intervention. We also compare dengue incidence in each of the treatment statuses with incidence figures in the control group. In both models, the unit of analysis is the census tract. We employ a differences-in-differences estimator to measure the treatment effects [31].

### 2.1 Ethics Statement

We pre-registered the experiment on the Evidence in Governance and Policy (EGAP) website (#20180504AA, https://osf.io/6q8vu/), and received IRB approval from New York University (IRB-FY2017-17) and Fundação Getulio Vargas (IRB-01/2017). The funders of this study had no influence over the formulation, organisation, analysis, and interpretation of the outcomes. Healthcare workers gave written consent upon filling the pre-treatment questionnaire. During the intervention, the occupants of households inspected for breeding sites and larvae gave verbal consent. For more information about the experimental design, please refer to the Supplementary Material.

## 3 Results

We find that financial incentives significantly increase the field productivity of healthcare workers fighting *Aedes aegypti*. Subjects in both the individual and the collective bonus groups locate more mosquito breeding sites and exterminate more larvae than participants in the control condition. Our results also show the intervention decreases city-wide levels of dengue incidence, but the coefficients only reach statistical significance when we combine the two treatments and contrast them with the control group. Overall, the findings suggest that monetary payments are an effective method of improving health worker performance, yet more research is needed to assess whether higher labour productivity leads to fewer dengue-related hospitalisations.

### 3.1 Field Productivity

Table 2 summarises the experimental results. Subjects who receive monetary incentives find and clean more breeding sites than those who earned a flat compensation fee. Workers in the individual treatment group find 25.118 (CI: [16.51, 33.726]) more breeding sites than the non-incentivised control group. The effect for those in the collective bonus group is 21.921 (CI: [15.235, 28.607]), slightly smaller than for the individual treatment. This finding suggests that the treatment does improve the performance of healthcare workers.

**Table 2:**
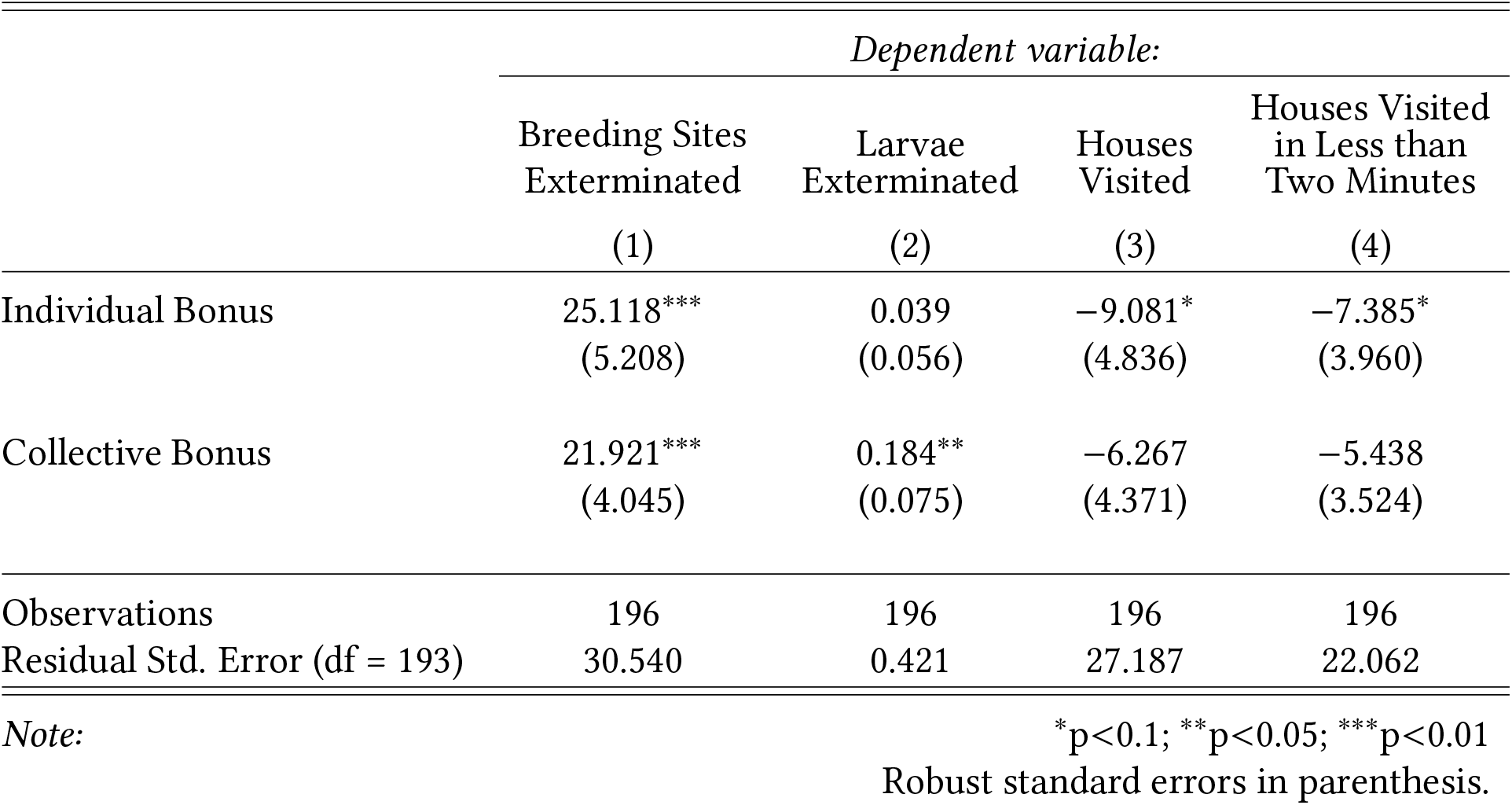
Field Productivity

|  | *Dependent variable:* |  |  |  |
| --- | --- | --- | --- | --- |
|  | Breeding Sites<br>Exterminated | Larvae<br>Exterminated | Houses<br>Visited | Houses Visited<br>in Less than<br>Two Minutes |
|  | (1) | (2) | (3) | (4) |
| Individual Bonus | 25.118***<br>(5.208) | 0.039<br>(0.056) | −9.081*<br>(4.836) | −7.385*<br>(3.960) |
| Collective Bonus | 21.921***<br>(4.045) | 0.184**<br>(0.075) | −6.267<br>(4.371) | −5.438<br>(3.524) |
| Observations | 196 | 196 | 196 | 196 |
| Residual Std. Error (df = 193) | 30.540 | 0.421 | 27.187 | 22.062 |

Note:
\*p<0.1; \*\*p<0.05; \*\*\*p<0.01 Robust standard errors in parenthesis.

Teams who received collective bonuses are the most effective in finding larvae. Subjects who worked under this treatment status have a 18.4% (CI: [6.1%, 30.7%]) higher likelihood of finding larvae than those in the control group. Locating mosquito larvae is a difficult task, thus the probability of successfully completing the task increases when subjects work in teams. Furthermore, our collective bonus scheme was not excludable within the teams, thus healthcare workers had an incentive to cooperate and receive higher rewards.

Subjects who received monetary incentives visited fewer houses than those who did not earn financial rewards. The result is statistically significant for the individual treatment group. While the finding seems counter-intuitive, it actually attests the effectiveness of the treatment. This is because we offered incentives for subjects to exterminate larvae and find breeding sites, not to visit a greater number of households. As a result, participants in the treatment groups did a thorough job in each household, which explains the lower number of visits during the allotted time. We find that subjects in the individual and in the collective bonus groups visited −9.081 (CI: [-17.073, −1.088]) and −6.267 (CI: [-13.491, 0.956]) households, respectively.

Our last indicator, the number of houses visited within less than two minutes, provides further evidence in favour of our argument. On average, participants in the individual bonus treatment arm visited −7.385 (CI: [-13.931, −0.84]) houses than the control, and those in the collective bonus group visited −5.438 (CI: [-11.263, 0.387]). Again, the result for the individual bonus group is statistically significant at the 10% level.

### 3.2 Disease Incidence

We employ two models to evaluate the effect of monetary incentives on dengue incidence. The first is a differences-in-differences estimation that tests whether the intervention *per se* had any effect on dengue hospitalisations in the city of Rio Verde. The second model compares dengue incidence in each of the treatment statuses with incidence figures in the control group. The unit of analysis is the census tract. Table 3 displays the results for our first model, in which we measure the impact of our intervention after 4, 8, 12, and 16 weeks. The last column shows how collective incentives compare with individual bonuses, with the latter considered as the control group.

**Table 3:**
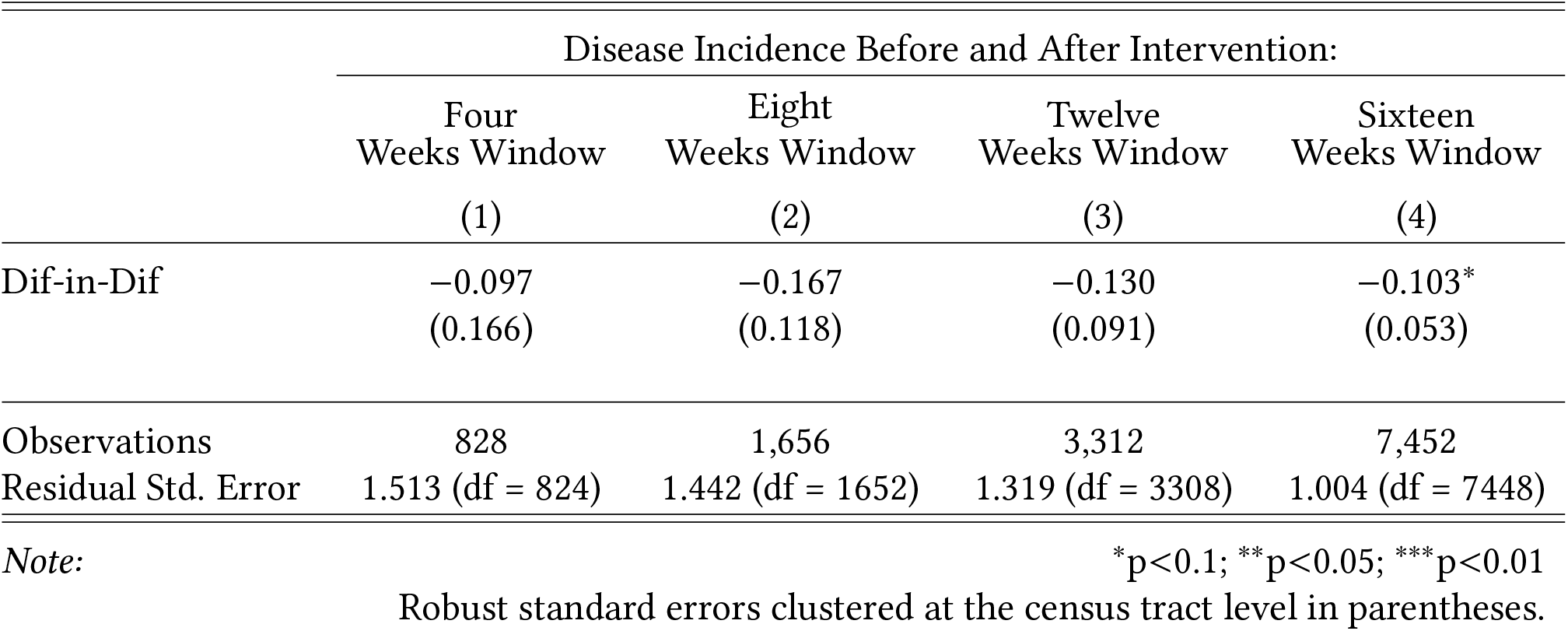
Differences-in-Differences Model

The intervention lowered dengue incidence in 10.3% (CI: [-18.9%, −1.6%]) if we take both treatments together and compare them with the control group. The result is consistent around fewer weeks before and after the intervention, but it does not reach conventional levels of statistical significance. This suggests that the treatment effect is probably small.

In our second model, we measure the effect of reward schemes on the number of hospitalisations per census tract. Table 4 shows hospitalisation counts before the intervention, and then 15, 30, 60, 90, and between 30 to 60 days after the treatment. Note that 96 pairs of healthcare workers analysed 139 census tracts. This is because 43 of those pairs worked in more than one census tract at once.

**Table 4:**
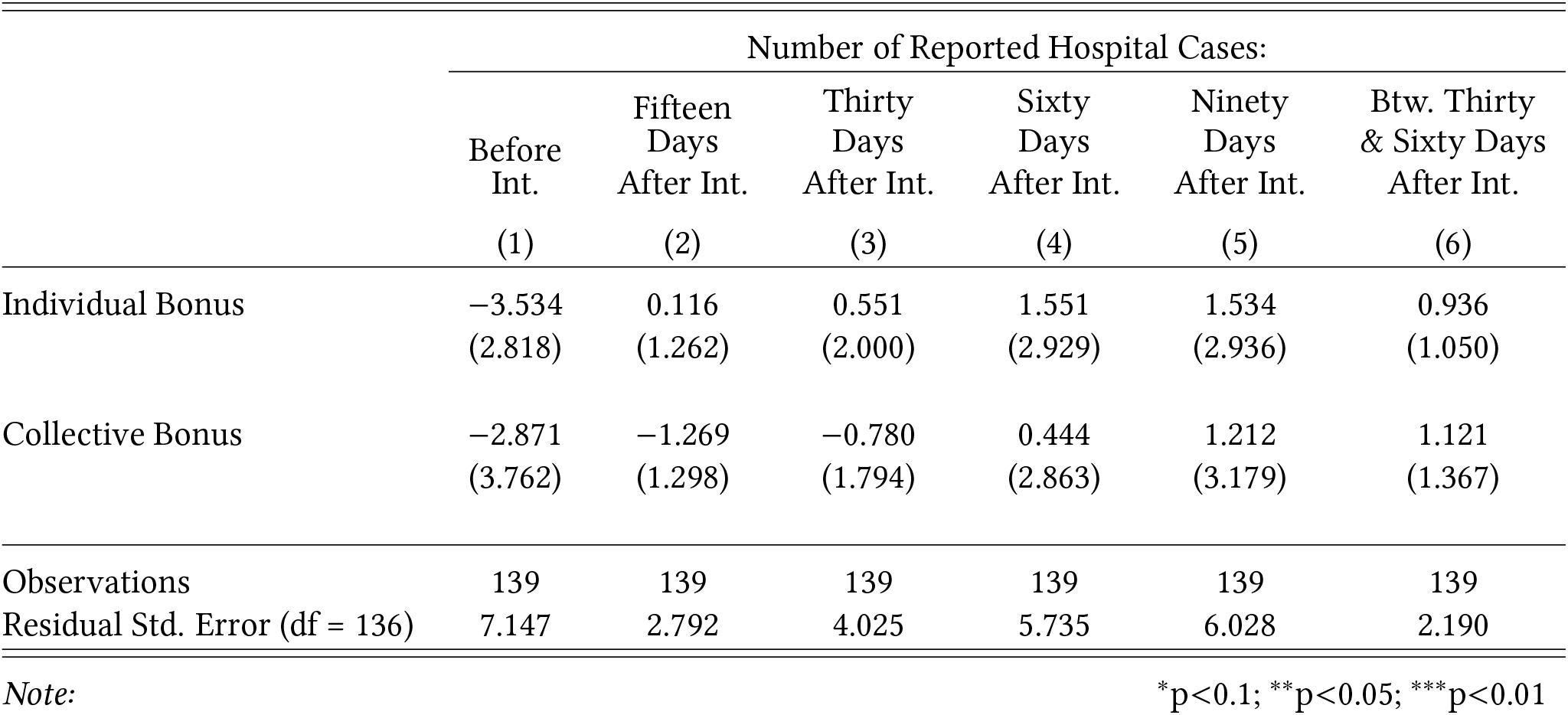
Disease Incidence

|  | Number of Reported Hospital Cases: |  |  |  |  |  |
| --- | --- | --- | --- | --- | --- | --- |
|  | Before<br>Int. | Fifteen<br>Days<br>After Int. | Thirty<br>Days<br>After Int. | Sixty<br>Days<br>After Int. | Ninety<br>Days<br>After Int. | Btw. Thirty<br>& Sixty Days<br>After Int. |
|  | (1) | (2) | (3) | (4) | (5) | (6) |
| Individual Bonus | -3.534<br>(2.818) | 0.116<br>(1.262) | 0.551<br>(2.000) | 1.551<br>(2.929) | 1.534<br>(2.936) | 0.936<br>(1.050) |
| Collective Bonus | -2.871<br>(3.762) | -1.269<br>(1.298) | -0.780<br>(1.794) | 0.444<br>(2.863) | 1.212<br>(3.179) | 1.121<br>(1.367) |
| Observations | 139 | 139 | 139 | 139 | 139 | 139 |
| Residual Std. Error (df = 136) | 7.147 | 2.792 | 4.025 | 5.735 | 6.028 | 2.190 |

Note:
\*p<0.1; \*\*p<0.05; \*\*\*p<0.01

We see that neither treatment has an impact on disease incidence in the Rio Verde census tracts. The treatment effect is not statistically different from zero in any model specification. Yet, the coefficients are negative, which indicates that had an impact occurred, the intervention would have lowered dengue incidence in the treated areas.

## 4 Discussion

We show that monetary incentives increase the productivity of community health care workers in Brazil. We contrast a control group (which received a flat performance fee) with two treatment groups, one which received an individual performance bonus, and second one that received a collective performance incentive. We find that both treatments increase the number of breeding sites encountered, and that collective incentives outperform individual performance rewards when the task is more laborious and excludable (exterminating larvae). Lastly, the control group presented a high incidence of houses visited in less than two minutes apart from each other, which suggests that the workers may not carry their tasks as intended in the absence of incentives. In a nutshell, monetary incentives work.

Our conclusions are consistent with results from comparable studies conducted in other settings, such as in education [32] and in private firms [33]. The findings also lend further evidence that incentivising teams is particularly effective when they carry out labour-intensive tasks, which opens a new avenue for research in preventive healthcare.

Regarding the effect of incentives on city-level dengue incidence, we find only mixed results. Why did the treatment not work as we planned? We believe the treatment was not as strong as it needed to be. Our treatment probably did not cover enough areas to ensure there was no mosquitos around the assigned blocks. As mosquitos can travel for long distances, the treatment might not have been sufficient. Also, Rio Verde had significant rates of dengue incidence during our experiment, reaching 3,411 reported infections from January to September 2018 [28]. Therefore, the treatment might have stronger effects only when applied for longer periods of time or in more neighbourhoods.

In this respect, our study suggests is that even though healthcare workers may be motivated, their efforts do not necessarily lead to a reduction in mosquito infections. Higher field productivity does not guarantee that the *Aedes aegypti* mosquito can be eradicated. To our knowledge, this issue has received little attention in the literature. Future research may explore the reasons why this is the case and if the disconnect between the two can also be identified in other areas.

### 4.1 Limitations

The study has several limitations. First, we ran our experiment for a small period of time and in a particular Brazilian city, thus the findings may not generalise to other locations. Second, our experiment needs a broader coverage to measure whether the treatment has a significant impact on dengue incidence, as the effect size is smaller than we expected. Third, we included a post-treatment adjustment to ensure the balance of our territorial variables. Although using matching estimators to improve balance is good statistical practice, the method requires additional assumptions. Finally, the experiment only measures the impact of financial rewards on worker performance, and it does not assess the effect of non-monetary incentives, such as motivational messages or constant feedback, which may be useful alternatives in jurisdictions where legal or financial constraints prevent the government from distributing financial bonuses to civil servants.

### 4.2 Public Health Implications

The experiment has important lessons for policy-making. *A. aegypti*-borne diseases are a significant public health concern, especially in developing nations. Results of this study indicate that monetary rewards play a major role in motivating civil servants, and that peer incentives are more cost-effective than individual bonuses. Finally, the article shows that anti-mosquito campaigns require a sustained effort to guarantee their success, and we still have no robust evidence that worker productivity alone is sufficient to reduce dengue infections.

## Data Availability

Replication materials are available at http://github.com/danilofreire/incentives-healthcare.

