## Supplementary material for "Financial Incentives and Healthcare Provision: Evidence from an Experimental *Aedes aegypti* Control Programme in Brazil": Online Appendix

Danilo Freire* Umberto Mignozzetti†

25 February 2021

*Independent Researcher,, http://danilofreire.github.io.

### A A Theory of Service Provision under Moral Hazard in Teams

Consider the following interaction between healthcare workers and a government agency.[1] The agency hires the workers to provide preventive healthcare. And the workers decide the amount of effort they will exert, conditional on the compensation scheme they receive.

Workers perform preventive healthcare tasks. To inspect a house and eliminate mosquito breeding sites, healthcare workers visit the households in pairs or in groups of three. Supposedly, this increases efficiency as it helps the team cover more territory and be more careful in the areas they inspect individually. Team structures also help in terms of lowering free-riding and increasing incentives for cooperative work. To capture this, we assume that a given worker $i$ divides her work effort into two tasks: her own effort ($e_i$) and helping efforts ($h_i$). To capture the costs of effort, we assume the following convex cost function $\frac{c}{2}(e_i^2 + h_i^2)$, where $c > 0$ represents the changes in marginal costs of effort and help.

Due to the nature of preventive care work, the policy outcome is probabilistic. Preventive healthcare aims to lower the chance of a person getting sick. For example, washing one's hands lowers the chance of flu infections, but from time to time a person gets the flu regardless of how many times she washes her hands. The only difference is that a person who washes her hands constantly has a lower chance of getting sick and will get sick fewer times during her lifespan. For this reason, we denote the probability of a successful provision of the service by a pair $i$ and $j$ of healthcare workers, as $P(e_i, h_i, e_j, h_j)$. For simplicity, we assume that the probabilities are separable, with $P(e_i, h_i, e_j, h_j) = P_i(e_i, h_j) + P_j(e_j, h_i)$. To make the output more concrete, assume that a given worker $i$ has an individual probability of $P_i(e_i, h_j) = e_i(1 + h_j)$ of successfully lowering the disease burden with her effort ($e_i$) and with the help of her peer ($h_j$).

From the perspective of the agency, successful work grants a benefit of $B > 0$. This benefit represents the social gains of lowering disease incidence in the municipality. For example, the payoff could be the amount of money saved from fewer hospitalisations or the economic benefit of fewer people missing work due to *A. aegypti*-transmitted diseases. The agency incentivises workers via three types of salaries: a flat salary without bonuses, an individual compensation, and a bonus when the entire team achieves their collective objectives. These formulations allow us to disentangle which scheme offers the highest yield in terms of disease prevention.

[1] This model was inspired by Bolton et al. (2005) and Itoh (1991). Here we present a particular case of their model to illustrate the effects of our treatment.

The optimal scheme to improve welfare is a contract where the government agency offers a compensation package, and the healthcare providers choose their effort accordingly. From the perspective of the worker, the expected utility of effort, provided that the compensation depends on successful provision of the service, and that the benefit from salary is given by a concave function $u(.)$. For concreteness, we use the function $u(x) = \sqrt{x}$. The expected utility of effort is then the compensation for each scenario, times their probability. It is equal to:

$$\begin{aligned} U_i(e_i, h_i; e_j, h_j) \quad = \quad & P_i(e_i, h_j)P_j(e_j, h_i)u(s + w_{both}) + \\ & P_i(e_i, h_j)(1 - P_j(e_j, h_i))u(s + w_{one}) + \\ & (1 - P_i(e_i, h_j))P_j(e_j, h_i)u(s + w_{one}) + \\ & (1 - P_i(e_i, h_j))(1 - P_j(e_j, h_i))u(s) - \frac{c}{2}(e_i^2 + h_i^2) \end{aligned}$$

And this utility is symmetric for the worker $j$. To simplify things, consider that the compensation under an unsuccessful service provision is equal to zero: $u(s) = 0$, or $s = 0$. From the perspective of the government agency, the benefit from offering the compensation is equal to the social benefit in each provision scenario minus the compensation times the likelihood of the scenario:

$$\begin{aligned} U_A(w_{both}, w_{one}, s; e_i, h_i, e_j, h_j) \quad = \quad & P_i(e_i, h_j)P_j(e_j, h_i)(2B - 2w_{both}) + \\ & P_i(e_i, h_j)(1 - P_j(e_j, h_i))(B - 2w_{one}) + \\ & (1 - P_i(e_i, h_j))P_j(e_j, h_i)(B - 2w_{one}) \end{aligned}$$

The situation is a standard contract theory problem, in which we have three compensation packages: no bonus ($w_{both} = w_{one} = 0$), individual bonus ($w_{both} = w_{one} \neq 0$), and collective bonus ($w_{both} \neq w_{one}$, $w_{both} \neq 0$, and $w_{one} \neq 0$). The optimal contract is then the solution for the following constrained optimisation problem:

$$\max_{w_{both}, w_{one}} U_A(w_{both}, w_{one}; e_i, h_i, e_j, h_j), \text{ subject to:}$$

$$(e_i, h_i) \in \arg\max U_i(e_i, h_i; e_j, h_j, w_{both}, w_{one})$$

$$(e_j, h_j) \in \arg\max U_j(e_j, h_j; e_i, h_i, w_{both}, w_{one})$$

And by choosing an optimal compensation scheme, the agency can induce more effort and more cooperation among workers.

#### A.1 Baseline: No Bonus

When the agency only pays a flat salary without performance incentives, the agency sets the performance compensation equal to zero: $w_{both} = w_{one} = 0$. This changes the expected utility of the agent to the following:

$$U_i(e_i, h_i; e_j, h_j) = -\frac{c}{2}(e_i^2 + h_i^2)$$

In this setting, exerting effort generates only costs and no benefits. The agents exert zero effort and help in equilibrium: $e_i^* = e_j^* = 0$ and $h_i^* = h_j^* = 0$. The agency pays the minimum wage accepted by law, and in this model, we assume that this wage is equal to zero. Accordingly, the agency's expected benefit is equal to:

$$R_A(\text{no bonus}) = 0$$

The prediction here is that workers will exert no effort, and that the agency will have only costs without accruing any benefit. This is the worst-case scenario in terms of the agency's welfare. In the experiment, it matches the control condition.[2]

[2]Throughout the article, we assume that the optimal output for the agency is equal to the maximum welfare for society. Although this is not always the case, it is a reasonable simplification for the purposes of this article. Adding a difference here would only complicate our results and provide no further insight into designing an incentive scheme to maximise welfare.

#### A.2 Performance Bonus for Individuals

When the benefits are tied to individual performance, the government agency compensates the workers for their effort, without tying their bonuses to helping other workers. This means that $h_i^* = h_j^* = 0$, but the optimal level of effort is always positive. Let $\alpha$ the bonus the government agency offers to workers conditional on lowering the incidence of the disease. Each worker then evaluates the utility of the bonus as $u_i(\alpha) = \sqrt{\alpha}$, as the utility of the agent is concave, representing the risk aversion of the agent. The optimisation problem, from the perspective of the agent, is to maximise her expected utility, given the bonus offered by the government agency upon achieving a successful provision:

$$\begin{aligned}\max_{e_i} U_i(e_i) &= \max_{e_i} \left[ P_i(e_i, 0)\sqrt{\alpha} - \frac{c}{2}(e_i^2) \right] \\ &= \max_{e_i} \left[ e_i\sqrt{\alpha} - \frac{c}{2}(e_i^2) \right]\end{aligned}$$

Both workers face the same incentive structure, so the equilibrium is symmetric, with $e_i = e_j$. Taking the derivative and equating it to zero, we find the following optimal effort level: $e_i^* = e_j^* = \frac{\sqrt{\alpha}}{c}$. The optimal level of effort represents the point where the marginal benefit from the government agency's compensation is equal to the marginal cost of effort. The benefit of a successful provision of the healthcare service is equal to – for both players – the total benefit times the chance of a successful provision times the difference between the benefit accrued from a successful provision minus the bonus paid for the worker. To compute the optimal contract the government agency has to offer in this setting, we must choose $\alpha$ to maximise the agency's expected utility:

$$\max_{\alpha} R_A(e_i^*(\alpha), e_j^*(\alpha)) = \max_{\alpha} \left[ 2\frac{\sqrt{\alpha}}{c}(B - \alpha) \right]$$

The optimal bonus is equal to $\alpha^* = \frac{B}{3}$. Plugging the optimal bonus and the optimal effort levels into the welfare function, we find the agency's expected revenue, which in this case is equal to:

$$R_A(\text{individual bonus}) = \frac{4}{c}\left[\frac{B}{3}\right]^{\frac{3}{2}}$$

The revenue is always greater than zero, provided that the costs of investing effort and the social benefits of the policy are greater than zero.

#### A.3 Performance Bonus for Teams

When the government agency sets rewards for both individual and team performance, healthcare workers divide their time between expending effort in their job ($e_i$) and helping their teams to get their jobs done ($h_i$). Helping may take many forms: from simple peer pressure and monitoring to ensure the peer is doing the job well, to efficiently dividing tasks and exploiting synergies in their duties. From the government agency's perspective, incentivising workers to help each other can increase the chances of obtaining successful service provision. From the standpoint of healthcare workers, optimal effort becomes a mix of her own efforts and the helping efforts that maximise her expected utility, provided that both efforts are incentive compatible. Incentive compatible means, in the case of healthcare workers, that both efforts are exerted to match the governmental agency's contractual incentives. The expected utility for the agent $i$ consists of the chance of getting a bonus times the utility of the bonus. This is iterated in each of the possible service provision scenarios.

$$\begin{aligned}\max_{(e_i,h_i)} U_i(e_i, h_i; e_j, h_j) &= \max_{(e_i,h_i)}\{e_j(1+h_i)e_i(1+h_j)u(w_{ij}) + \\ &\quad e_j(1+h_i)(1-e_i(1+h_j))u(w_j) + \\ &\quad e_i(1+h_j)(1-e_j(1+h_i))u(w_i) - c\frac{e_i^2+h_i^2}{2}\}\end{aligned}$$

Again, both players are *ex-ante* similar in terms of their expected utilities. This allows us to solve for the Symmetric Nash Equilibrium, where:

1. The effort of a given healthcare provider ($i$) is equal to the effort of the other healthcare provider ($j$): $e_i = e_j = e$.
2. The help of one healthcare provider is equal to the help of the other healthcare provider $h_i = h_j = h$.
3. Both agents have the same utility function, $U_i(.) = U_j(.)$.

Maximising the expected utility leads us to the optimal effort and amount of help for the healthcare provider, given the contractual scheme. The optimal provision must satisfy the following equation:

$$\frac{\partial U}{\partial e} = e(1+h)^2(u(w_{ij}) - u(w_i) - u(w_j)) + (1+h)u(w_i) - ce$$
$$\frac{\partial U}{\partial h} = e^2(1+h)(u(w_{ij}) - u(w_i) - u(w_j)) + eu(w_j) - ch$$

To simplify our computation, we consider contracts that vary in one dimension $\alpha$. The individual part of the contractual benefit is equal to multiples of this contract for each situation: own successful provision and the successful colleague provision.

In this example, we restrict our attention to contracts where $u(w_i) = u(w_j) = \sqrt{\alpha}$ and $u(w_{ij}) = 2u(w_i)$. These contracts are a better match for the conditions we set in the experiment: they represent the collective gain when recompensing the agent for peer and individual successes. The optimal effort and help become:

$$e_i^* = e_j^* = \frac{\sqrt{\alpha c}}{c - \alpha}$$

and

$$h_i^* = h_j^* = \frac{\alpha}{c - \alpha}$$

The governmental agency now maximises the bonus for the individual provision ($\alpha$), given that the collective provision is higher than it is. The expected revenue of the agency is equal to the chance of lowering the disease burden times the benefit minus the costs of paying the salaries of the healthcare providers:

$$\max_{\alpha} R_A(e_i^*(\alpha), h_i^*(\alpha), e_j^*(\alpha), h_i^*(\alpha)) = \max_{\alpha} \left[ 2\frac{c\sqrt{c\alpha}}{(c-\alpha)^2}(B - 2\alpha) \right]$$

The derivative of this equation is the polynomial in the second degree in $\alpha$. By the intermediate value theorem, it is easy to see that it has one solution for $\alpha \in [0, B]$. To illustrate, consider Figure 1, where we plot the expected revenue for the agency ($R_A(\alpha)$), as a function of the offered compensation $\alpha$, and the three types of contracts: no productivity incentives, individual productivity incentives, and team productivity incentives, fixing $B = c = 1$.

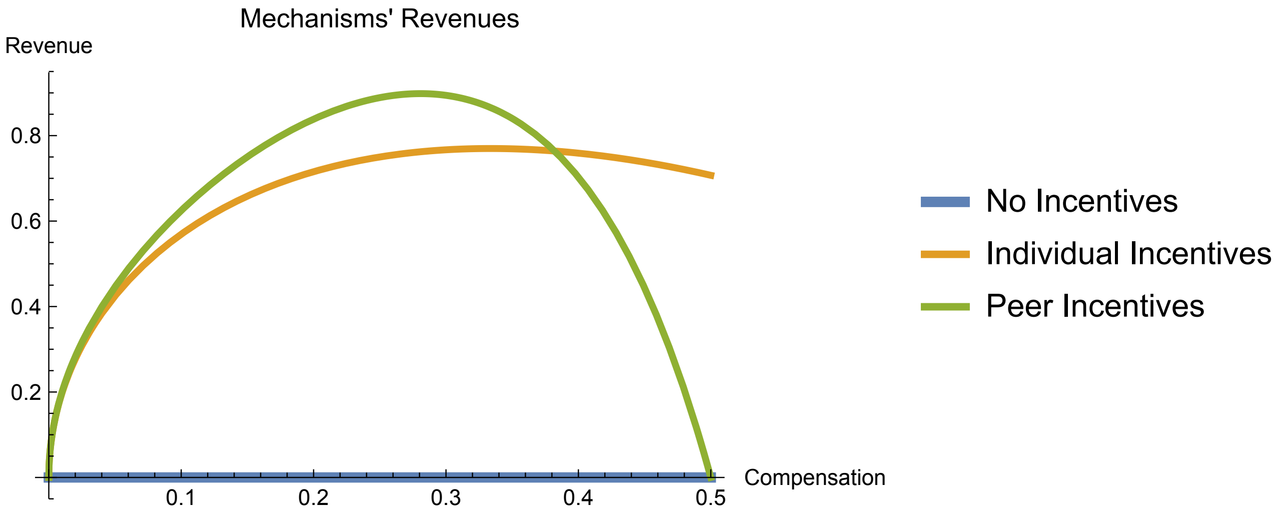

Figure 1: Governmental Agency's Revenue in Terms of Contract and Compensation

Therefore, the maximum benefit that the agency can achieve is higher when the agency compensates the workers for their cooperation. When the agency incentivises team effort, workers can do better, on margin, by sharing the benefit of successful provision.[3]

### B Data Analysis

#### B.1 Data Sources

We use five data sources in this study. All of them are included in `../data/incentives_healthcare.RData`:

- `pretreat`: Pre-treatment balance information.
- `prodfr`: Field productivity data.
- `dfr`: Disease incidence in every Rio Verde census sector.
- `dfr2`: Disease incidence in every Rio Verde census sector that received the intervention.
- `did_full`: Differences-in-differences estimation that measures the effect of our intervention against a pure control group.

[3]The limits for this reasoning appear when there are gains of specialisation. For instance, if the costs for effort and help were such that $\frac{c(e+h)^2}{2}$, then diverting energy for help is costly, on margin, at the same rate as the given effort provision. In this case, the compensation to stimulate cooperation has to be significantly higher, and even in this case there will be incentives for not cooperating.

##### B.1.1 Pre-Treatment Balance Dataset

The pre-treatment balance dataset contains information about every individual who interacted with the research team at any point of time. To be included in this dataset, the person needs to have filled the Google Form we circulated on Facebook as part of our recruitment campaign. This dataset contains 13 variables that we collected before the intervention. We gathered these data either via Google Forms, or in pre-treatment surveys before the training session.

1. `Age`: Age of the participant.
2. `Female`: Dummy for female participant.
3. `HasVehicle`: Dummy for having a vehicle (car or motorcycle).
4. `aboveMedianAltr`: Dummy for ranking above the sample median in our altruism scale.
5. `relig`: Dummy for ranking above the sample median in our religiosity scale.
6. `polengage`: Dummy for ranking above sample median in political engagement.
7. `socengage`: Dummy for ranking above the sample median in social engagement.
8. `collegeplus`: Dummy for having college degree or higher.
9. `aboveTwoMinWageFam`: Dummy for earning more than two Brazilian minimum wages (R$ 900.00 per month).
10. `FBpop`: Dummy for having above median Facebook popularity.
11. `training`: Dummy for having joined the training session.
12. `participating`: Dummy for having participated in the field experiment.
13. `assignment`: Dummy for treatment status assignment.

##### B.1.2 Field Productivity Dataset

We collected field productivity data based on the subjects' performance during the experiment. We measured productivity with three indicators:

1. Number of houses visited.
2. Number of possible breeding sites exterminated.
3. Number of larvae exterminated.

We compiled these data using photos taken during the field experiment. All participants were instructed to take pictures of clean breeding sites, and to make videos showing the larvae they exterminated. After finishing the inspection of each household, subjects had to take a last picture of the house to prove that they had been there.

The final dataset has the following variables:

1. `D`: Treatment assignment.
2. `pair_n`: Pair index ID.
3. `nHouse`: Number of houses visited.
4. `nBreed`: Number of breeding sites cleaned.
5. `nLarvae`: Number of larvae exterminated.
6. `nHouseVisit`: Number of houses that were indeed visited (based on pictures of breeding sites and larvae videos).
7. `nHouseLessTwoMin`: Number of houses visited in less than two minutes (which denotes either that the pair did not go to the place, or that there were no residents at home).

##### B.1.3 Disease Incidence Datasets

To assess whether the intervention had an impact on the municipal healthcare system, we collected 13 variables in two distinct datasets. All datasets compile data from the National Health Notification System (SINAN) for the Rio Verde municipality. The dataset has the complete records of all individuals who had been hospitalised between four months before and four month after the intervention. We geolocated the household addresses of those individuals who were hospitalised with dengue fever, then we matched the case to the census sector where that person lives. Finally, we computed the number of cases before and after a few time windows around the intervention. The result is two datasets, `dfr` (with all census sectors) and `dfr2` (with only the census sectors matched in the intervention). Both datasets have the following variables:

1. `CensusSectors`: Last three digits of census sector (2010 Brazilian census).
2. `NHouses`: Number of houses in each census sector (2010 Brazilian census).
3. `NHouseholds`: Average Number of households in each house/building and in each census sector (2010 Brazilian Census).
4. `AvgHouseholdSize`: Average household income per census sector (2010 Brazilian Census).
5. `logAvgIncomeHousehold`: Log of average household income per census sector (2010 Brazilian Census).

6. D1: Treatment assignment.
7. D2: Intervention x no intervention (no intervention missing in dfr2).
8. nc15dafter: Number of cases 15 days after the intervention.
9. nc30dafter: Number of cases 30 days after the intervention.
10. nc60dafter: Number of cases 60 days after the intervention.
11. nc90dafter: Number of cases 90 days after the intervention.
12. nc30to60dafter: Number of cases between 30 and 60 days after the intervention.
13. plac: Number of cases before the intervention.

##### B.1.4 Differences-in-Differences Dataset

We build the did_full dataset to investigate whether our intervention had any impact on public health outcomes. The disease incidence data describe the treatment effect. In the differences-in-differences models, we combine all treatments and investigate how they perform against a pure control group, which are the census sectors that were not visited during the intervention.

The dataset has ten variables:

1. CensusSectors: Last three digits of census sector (2010 Brazilian Census).
2. D: Treatment status census sector (intervention versus pure control).
3. weekn: Week name.
4. ncas: Number of cases in a given week and census sector.
5. Tm: Week number.
6. Tm2: Intervention timing indicator.
7. DID: Differences-in-differences ($Tm2 \times D$).
8. ncas4weekwindow: Number of cases four weeks window around the intervention.
9. ncas8weekwindow: Number of cases eight weeks window around the intervention.
10. ncas12weekwindow: Number of cases twelve weeks window around the intervention.

#### B.2 Covariate Balance

We measure covariate balance using four statistics. First, we check whether subjects decided to take part in the training sessions. Second, we evaluate the balance among those who decided to participate in the experiment. Subjects had to take part in the training process and participate in the fieldwork.

Third, we check if individual characteristics are balanced across our treatment groups. As we mentioned above, we collected that information via Google Forms and using a pre-treatment survey. Finally, we check the territorial assignment to make sure that the pre-treatment characteristics are balanced across the places that subjects visited. We find good balance for the individual assignment (models 1 to 3), and we corrected eventual imbalances in the territorial assignment using propensity score matching (Rosenbaum and Rubin 1985).

##### B.2.1 Training Participation

Table 1: Covariate Balance on Training

| Variable | Not Training | Training | F-Statistic | P-Value |
|---|---|---|---|---|
| Age | 26.6 | 25.81 | 1.343 | 0.247 |
| Female | 0.74 | 0.74 | 0 | 0.99 |
| Has Vehicle? | 0.37 | 0.35 | 0.252 | 0.616 |
| N | 243 | 216 | NA | NA |

##### B.2.2 Participation in the Field

Table 2: Covariate Balance on Participation

| Variable | Not Participating | Participating | F-Statistic | P-Value |
|---|---|---|---|---|
| Age | 26.62 | 25.7 | 1.735 | 0.188 |
| Female | 0.73 | 0.74 | 0.02 | 0.886 |
| Has Vehicle? | 0.37 | 0.35 | 0.205 | 0.651 |
| N | 263 | 196 | NA | NA |

##### B.2.3 Individual Assignment

Table 3: Covariate Balance on Individual Characteristics

| Variable | Control | Individual | Collective | F-Statistic | P-Value |
|---|---|---|---|---|---|
| Age | 26.03 | 26.24 | 25.1 | 0.49 | 0.613 |
| Female | 0.71 | 0.71 | 0.79 | 0.862 | 0.424 |
| Has Vehicle? | 0.4 | 0.34 | 0.31 | 0.639 | 0.529 |
| Altruist | 0.59 | 0.64 | 0.65 | 0.321 | 0.725 |
| Religiosity | 0.42 | 0.37 | 0.51 | 1.476 | 0.231 |
| Pol. Engagement | 0.68 | 0.63 | 0.68 | 0.257 | 0.774 |
| Soc. Engagement | 0.63 | 0.6 | 0.57 | 0.225 | 0.799 |
| Above Two Min. Wage | 0.27 | 0.36 | 0.34 | 0.731 | 0.483 |
| Facebook Popularity | 0.49 | 0.57 | 0.51 | 0.534 | 0.587 |
| N | 78 | 70 | 68 | NA | NA |

##### B.2.4 Unweigted Territorial Assignment

Table 4: Covariate Balance Un-Weighted Territorial Assignment

| Variable | Control | Individual | Collective | F-Statistic | P-Value |
|---|---|---|---|---|---|
| Number of Houses | 276.41 | 243.58 | 255.2 | 2.368 | 0.098 |
| Number of Households | 885.67 | 750.34 | 785.18 | 3.765 | 0.026 |
| Avg. Household Size | 3.21 | 3.07 | 3.07 | 6.566 | 0.002 |
| Log Avg. Income | 6.47 | 6.86 | 6.85 | 9.213 | 0 |
| Cases Before Treatment | 13.78 | 5.94 | 4.66 | 15.638 | 0 |

##### B.2.5 Propensity-Score Weighted Territorial Assignment

Table 5: Covariate Balance Weighted Territorial Assignment

| Variable | No Bonus | Individual | Collective | F-Statistic | P-Value |
|---|---|---|---|---|---|
| Number of Houses | 276.41 | 282.04 | 283.5 | 0.178 | 0.837 |
| Number of Households | 885.67 | 903.04 | 907.14 | 0.154 | 0.858 |
| Avg. Household Size | 3.21 | 3.21 | 3.2 | 0.019 | 0.981 |
| Log Avg. Income | 6.47 | 6.45 | 6.46 | 0.071 | 0.931 |
| Cases Before Treatment | 13.78 | 10.24 | 10.91 | 1.791 | 0.171 |

#### B.3 Productivity Outcomes

Table 6: Field Productivity

| | *Dependent variable:* | | | |
|---|---|---|---|---|
| | Houses Visited | Houses Vis. in Less than Two Min. | Breeding Sites Terminated | Larvae Terminated |
| | (1) | (2) | (3) | (4) |
| Individual Bonus | $-9.081^{*}$ | $-7.385^{*}$ | $25.118^{***}$ | 0.039 |
| | (4.836) | (3.960) | (5.208) | (0.056) |
| Collective Bonus | $-6.267$ | $-5.438$ | $21.921^{***}$ | $0.184^{**}$ |
| | (4.371) | (3.524) | (4.045) | (0.075) |
| Observations | 196 | 196 | 196 | 196 |
| Residual Std. Error (df = 193) | 27.187 | 22.062 | 30.540 | 0.421 |
| *Note:* | | | | $^{*}$p<0.1; $^{**}$p<0.05; $^{***}$p<0.01 |

#### B.4 Disease Incidence Outcomes

Table 7 shows that no treatment effect reaches conventional levels of statistical significance.

Table 7: Disease Incidence

| | Number of Reported Hospital Cases: | | | | | |
|---|---|---|---|---|---|---|
| | Before Int. | Fifteen D. After Int. | Thirty D. After Int. | Sixty D. After Int. | Ninety D. After Int. | Btw. Thirty & Sixty D. After Int. |
| | (1) | (2) | (3) | (4) | (5) | (6) |
| Individual Bonus | $-3.534$ | 0.116 | 0.551 | 1.551 | 1.534 | 0.936 |
| | (2.818) | (1.262) | (2.000) | (2.929) | (2.936) | (1.050) |
| Collective Bonus | $-2.871$ | $-1.269$ | $-0.780$ | 0.444 | 1.212 | 1.121 |
| | (3.762) | (1.298) | (1.794) | (2.863) | (3.179) | (1.367) |
| Observations | 139 | 139 | 139 | 139 | 139 | 139 |
| Residual Std. Error (df = 136) | 7.147 | 2.792 | 4.025 | 5.735 | 6.028 | 2.190 |
| *Note:* | | | | | | $^{*}$p<0.1; $^{**}$p<0.05; $^{***}$p<0.01 |

#### B.5 Differences-in-Differences Estimation for Healthcare Outcomes

The results for the differences-in-differences estimations are available in Table 8.

Table 8: Differences-in-Differences Model

| | Disease Incidence Before and After Intervention: | | | |
|---|---|---|---|---|
| | Four Weeks Window | Eight Weeks Window | Twelve Weeks Window | Sixteen Weeks Window |
| | (1) | (2) | (3) | (4) |
| Dif-in-Dif | −0.097 | −0.167 | −0.130 | $-0.103^{*}$ |
| | (0.166) | (0.118) | (0.091) | (0.053) |
| Observations | 828 | 1,656 | 3,312 | 7,452 |
| Residual Std. Error | 1.513 (df = 824) | 1.442 (df = 1652) | 1.319 (df = 3308) | 1.004 (df = 7448) |
| *Note:* | | | | $^{*}$p<0.1; $^{**}$p<0.05; $^{***}$p<0.01 |

The results suggest the following:

1. There is a 10.3% effect on the full data model.
2. The coefficient for the other windows are not significant but the signs are consistent with lower disease incidence.

### C Experiment Report

Here we follow the American Political Science Association Experimental Session Report Guidelines (Gerber et al. 2014) and present a detailed discussion of our sample and research design.

#### C.1 Hypothesis

##### C.1.1 Question Addressed by the Experiment

We study the impact of monetary incentives on the performance of healthcare workers in Brazil. Our goal is to find which financial incentive scheme improves healthcare worker productivity the most.

Several articles highlight that higher payments improve public service provision (e.g., Doran et al. 2017; Duflo et al. 2012; Langdown and Peckham 2014). However, these papers focus on services that are provided individually, such as by a single a teacher in a classroom. In public health, it is rare that employees work independently. This, in turn, poses the question: what is the best way to improve public healthcare provision, when the services are delivered by teams rather than by individuals?

Here we try to bridge this gap by testing whether team incentives are more effective than individual incentives to improve preventive healthcare provision in Brazil.

The Brazilian case is well suited for this test for two main reasons. First, *A. aegypti*-borne diseases are some of the leading causes of congenital disabilities, hospitalisation, and casualties in the Global South. Second, Brazilian healthcare agents work in pairs when they are allocated to visit households and exterminate mosquito breeding sites. Therefore, we could test individual versus team bonuses in a setting that is relevant to welfare.

##### C.1.2 Hypothesis Tested

We tested which types of monetary bonus improves the performance of healthcare workers in Brazil. We tested three incentive schemes: first, monetary compensation with no performance bonuses; second, individual compensation for performances above the median of the headquarter they were assigned; and third, peer bonus compensation for team performances above the median of their headquarters. Theoretically, team bonuses should be more effective, as they increase peer monitoring and reduce the free-rider problem. However, if there are few gains from synergies among workers, individual rewards can be more useful.

#### C.2 Subjects and Context

##### C.2.1 Why Was This Subject Pool Selected?

We selected participants via Facebook recruitment advertisements, which we posted to citizens of the Municipality of Rio Verde, Goiás State, Brazil. We chose that city because it had a considerable spike in dengue fever cases in 2017 and 2008, what lead us to establish a partnership with the Mayor's Office especially to run the experiment we describe here.

We sent an email to participants and asked a few questions on demographics and other characteristics.

##### C.2.2 Who Was Eligible to Participate in the Study?

Adults who were 18 years of age or older.

##### C.2.3 What Would Result in the Exclusion of a Participant?

Being under-age or not participating in the training lessons or the intervention.

##### C.2.4 Were Any Aspects of Recruitment Changed after the Recruitment Began?

No.

##### C.2.5 Procedures Used to Recruit and Select Participants

We posted a Facebook ad in the municipality participating in the intervention. The ad invited potential participants to join the research and explained the payment scheme. If they clicked on the ad, they were directed to a Google Form, where they added their email address and contact information.

##### C.2.6 Recruitment Dates

The online recruitment took place right after respondents filled up the form. We sent a link containing a pre-treatment survey and some information about the *A. aegypti* mosquito to potential participants. After respondents completed the survey, we informed them about the in-person training dates and asked them to select their preferred times to participate. They had to join us for a two-hour training session.

The training date for the pilot was the $13^{\text{th}}$ of April, 2018. The training date for the intervention was $4^{\text{th}}$ of May, 2018.

##### C.2.7 Settings and Locations Where the Data Were Collected

We use two primary datasets in the paper. First, we collected productivity data from the cell phones used by the participants in the field. Second, we collected governmental data on *A. aegypti*-borne diseases from the National Health Notification System (SINAN).

The first dataset was generated by participants themselves and included the pictures taken during their fieldwork. The second dataset is comprised of governmental records of individuals that checked into hospitals and diagnosed with dengue fever. These data include the individual's home address, which we used to geolocate his or her household. Among the diseases transmitted by *A. aegypti*, dengue fever is the most prevalent and the easiest one to detect.

##### C.2.8 Relevant Information about the Population

All participants were from the municipality of Rio Verde, in the state of Goiás, Brazil. A map of the municipality follows below:

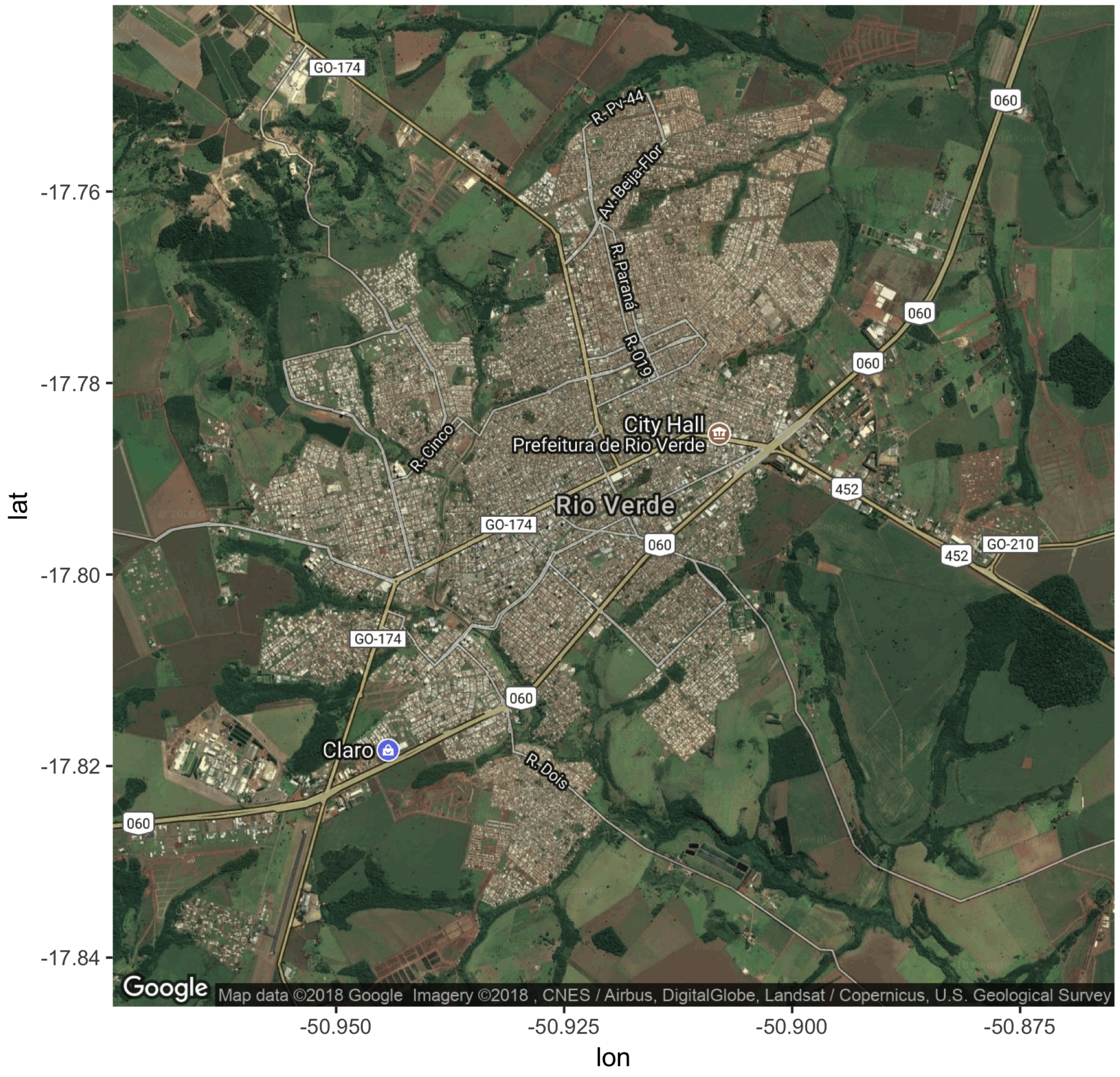

Figure 2: Map of Rio Verde, GO, Brazil.

#### C.3 Allocation Methods

##### C.3.1 Randomisation procedure

We generated the treatment assignment using block randomisation applied on seven variables:

1. Age
2. Altruism.
3. Religiosity score.
4. Political engagement.
5. Social engagement.
6. Family income higher than two minimum wages.
7. Facebook popularity (number of friends).

We computed these variables from our pre-treatment survey and Google Forms. To test the covariate balance, we used F-tests on the block randomisation data. The balance tests follow in the data analysis section.

#### C.4 Random Assignment

##### C.4.1 Units of Randomisation

We randomised the treatment at the participant level.

##### C.4.2 Cluster Random Assignment

We did not employ cluster random assignment. However, in the differences-in-differences estimation, we use the census sector where the pair worked to cluster the standard errors.

#### C.5 Blinding

##### C.5.1 Were Participants Unaware of the Treatment Assignment?

Yes. They knew about their incentive schemes, but not about the other incentive schemes we implemented in our experiment. To ensure that there would be no spillovers, we divided each treatment assigned teams into three different headquarters:

1. Assembleia de Deus Church (No bonus)
2. Videiras Church (Individual reward)

3. Sara Nossa Terra Church (Collective bonus)

These headquarters were away from each other to avoid contact between participants.

##### C.5.2 Were Those Administering the Intervention Unaware of the Random Assignment?

No. The head research assistants in each group knew that different treatment would be administered in other research headquarters. We took this measure because in case of treatment spillover we would have to quickly remove the pairs who knew about the treatment conditions.

##### C.5.3 Checked Whether Blind was Successful?

The single-blind was successful. We received no complaints during the intervention day. After the intervention day, participants became aware of the experiment. The post-treatment spillover made the post-treatment survey ineffective because they refused to cooperate with the researchers.

#### C.6 Treatments

##### C.6.1 Treatment Groups

We have one control and two treatment groups in our experiment:

1. Control (no performance bonus)
2. Individual treatment
3. Collective treatment

In the individual treatment group, we explained to participants that we would rank their individual performance based on number of visited houses, number of breeding sites removed and cleaned, and number of larvae exterminated. Participants would have their performance ranked and the ones above the median would double their monetary reward (from BRL 110.00 to BRL 220.00).

In the collective treatment group, we informed participants that we would rank their team performance based on number of visited houses, number of breeding sites removed and cleaned, and number of larvae exterminated. We would sum their individual performance with the performance of their field peers, and then rank the teams. Teams with performance above the median would double their monetary reward (from BRL 110.00 to BRL 220.00 for each person in the team).

The fieldwork was performed in pairs, to mimic the implementation of existing mosquito control strategies in the area.

##### C.6.2 Control Group

The control group received a monetary reward with no performance requirement.

##### C.6.3 Experimental Instructions

At 5:00 on intervention day, we randomly assigned pairs and sent emails to the individuals directing them to each of the headquarters, based on their treatment status.

We had three headquarters: the headquarters for the control group (Assembleia de Deus church); the headquarters for the individual treatment group (Videiras church); and the headquarters for the peer treatment group (Sara Nossa Terra church). We printed set of leaflets with places that they had to visit in order to administer the treatment. One example leaflet follows below:

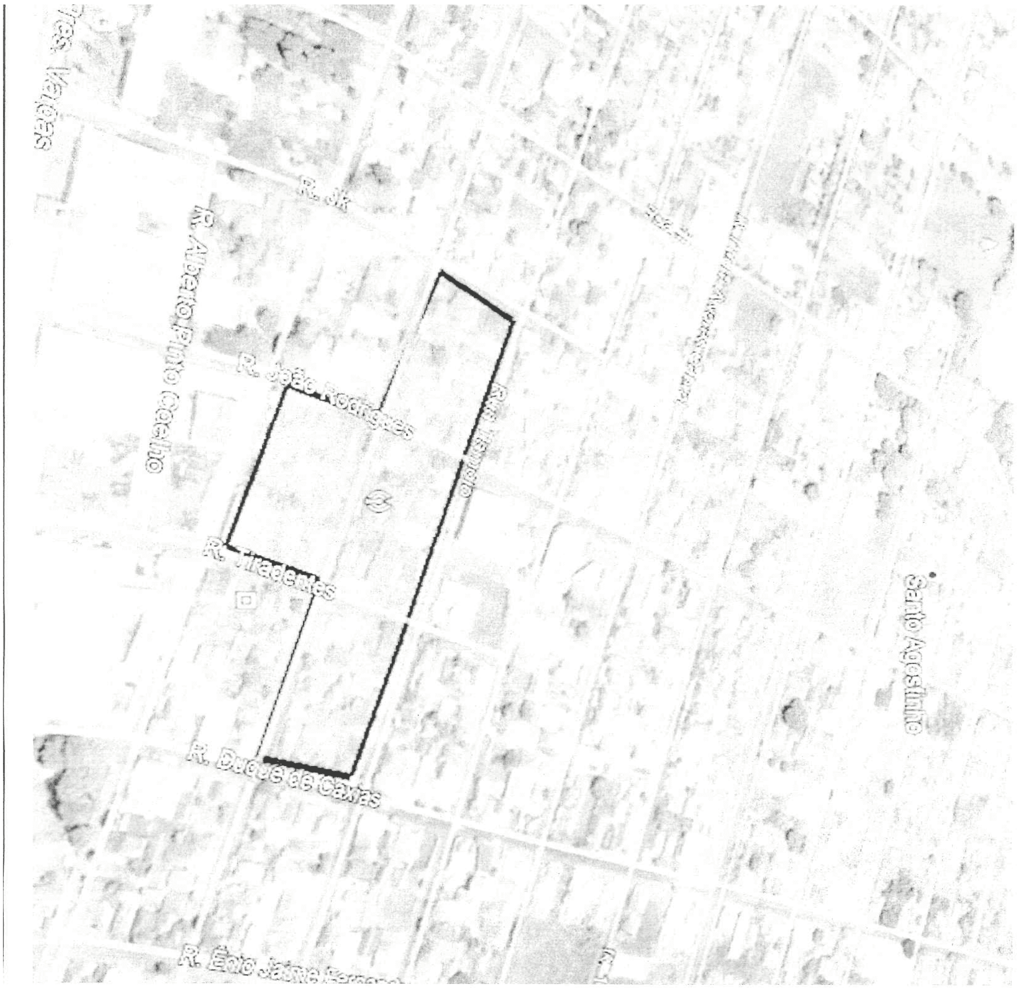

Figure 3: Example Leaflet.

And the working areas follow below:

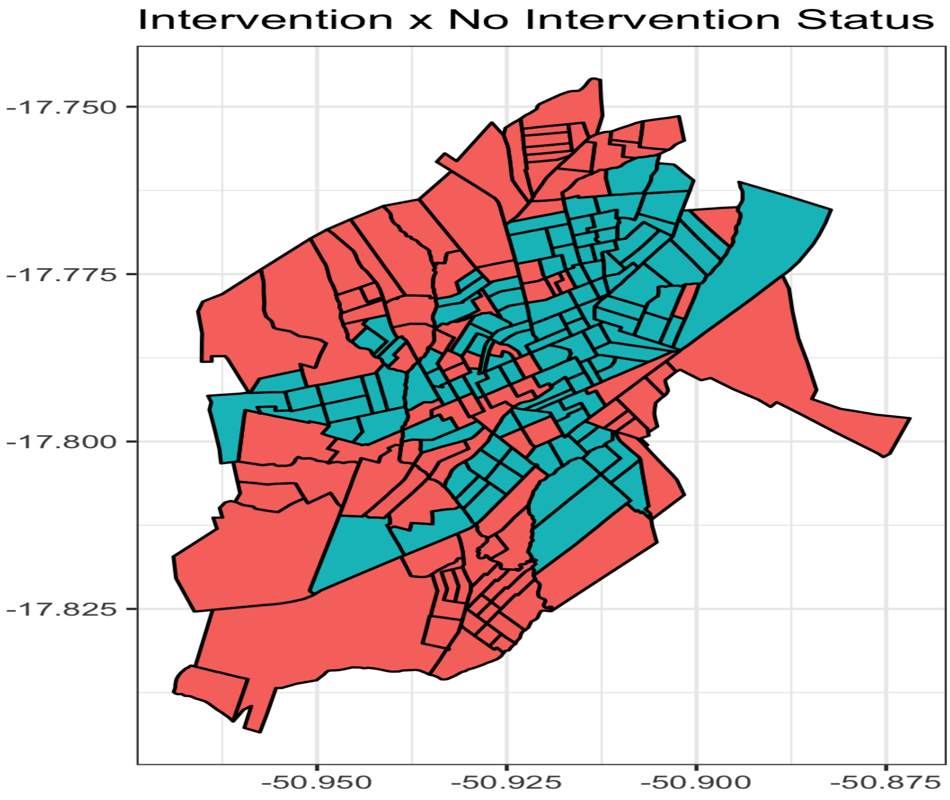

Figure 4: Example Working Area.

We administered the main treatments at 11:00. They consisted in telling the following instructions to participants after we handed the leaflet with their working places:

- **Control:** we told them that there would be an increase in their payment without explaining the reason. We told them about how their performance would be measured, without saying anything about compensation based on performance.
- **Individual treatment:** we told participants that we would measure their performances in the field, and that we would rank all performances. The individuals with performance above the median would receive the bonus. We also explained in lay terms what a median is and how we would measure their performance.
- **Peer treatment:** we told participants that we would measure their performances in the field, and that we would rank all performances. The pairs with performance above the median would receive the bonus. We also explained in lay terms what a median is and how we would measure their performance.

#### C.7 How and When Manipulations Were Administered

##### C.7.1 Delivery Methods

The manipulations were delivered in person by the chief headquarter RA. First, the manipulations was administered to all pairs simultaneously, then reinforced to each particular pair.

##### C.7.2 Software Used to Administer the Treatment

We used R to perform the randomisation process. The treatment administration was done in person.

#### C.8 Results

##### C.8.1 Outcome Measures

There are two groups of outcomes. First, the performance outcomes that we collected from the cellphone used by each participant in the field. Second, dengue incidence figures reported by the healthcare facilities in the municipality.

For the performance data we have:

1. Geolocated pictures of house fronts as a proof that they visited to house.
2. Pictures of exterminated breeding sites.
3. Video of exterminated *A. aegypti* larvae.

For governmental data, we have the total number of hospitalised individuals who were infected with dengue fever. There were no reported cases of Zika and Chikungunya.

##### C.8.2 Covariates

We collected demographic information from participants.

##### C.8.3 Which Outcomes and Subgroup Analysis Were Specified Prior to the Experiment?

The Rio Verde Mayor's Office did not hand in the infections data prior to the intervention. Therefore, we collected information on the neighbourhoods that had higher dengue fever incidence prior to the intervention, and implemented the territorial assignment accordingly. However, due to absence of micro-level data on infectious diseases, we did not consider them prior to the intervention.

##### C.8.4 Exploratory Analysis

We run a pilot study with 20 pairs before the main intervention. We did not administer the incentivised intervention in the pilot study to avoid contamination in the results. The pilot study helped us update our expectations regarding the productivity in the field. Previously, we projected that we should assign around 20 houses per pairs. However, in the pilot study we learned that pairs could visit from 40 to 60 houses per pair. In one-third of the households nobody answered the door, and in most of them the yards were too small to find any breeding sites. Moreover, most of the places with dengue in the previous week had been visited by the municipal health care workers.

#### C.9 CONSORT

##### C.9.1 Number of Subjects Initially Accessed for Eligibility

459 individuals filled in the Google Form. `sum(pretreat$training=='Training')` showed for the training session one day before the intervention. 196 individuals participated in the intervention and remained until the end of it.

##### C.9.2 Exclusions Prior to Random Assignment

From all eligible people, we excluded the ones who missed the training and the field implementation sessions. This accounts for 57.3% of the total.

##### C.9.3 Subjects Initially Assigned to Each Experimental Group

1. 68 to control group.
2. 70 to individual treatment group.
3. 78 to peer treatment group.

##### C.9.4 Proportion Received x Not Received Intervention

1. 58 received the control intervention out of 68 in the control group.
2. 64 received the intervention out of 70 in the individual treatment group.
3. 74 received the intervention out of 78 in the peer treatment group.

##### C.9.5 Why Did Subjects Not Receive Intervention?

The individuals did not receive the intervention if they missed the fieldwork date. They were excluded from the data.

##### C.9.6 Number Subjects Each Group Dropped Experiment

No subject were dropped from the assignment. The attrition was caused only by the fact that some individuals did not show up for the training and the treatment.

#### C.10 Statistical Analysis

##### C.10.1 Describe Statistical Analysis

We run four types of statistical analysis:

1. Balance tests.
2. Performance outcome analysis.
3. Disease incidence analysis.
4. Differences-in-differences analysis.

The main equations for the performance outcomes are:

$$Y_i = \alpha + \beta D_i + \varepsilon_i$$

Where $\beta$ is the outcome of interest, $Y_i$ is the outcome for case $i$, and $D_i$ is the treatment status for case $i$. And the code is:

```
mod <- lm(outc ~ treat, data = dataframe)
```

The main equations for the disease incidence are:

$$Y_i = \alpha + \beta D_i + \varepsilon_i$$

Where $\beta$ is the outcome of interest, $Y_i$ is the outcome for case $i$, and $D_i$ is the treatment status for case $i$. The only difference is that the estimates are weighted using the propensity score matching. And the code is:

```
mod <- lm(outc ~ treat,
          data = dataframe,
          weights = dataframe$weights)
```

Finally, the differences-in-differences estimator has the following form:

$$Y_i = \alpha + \gamma Time_i + \theta Treat_i + \beta Time_i \times Treat_i + \varepsilon_i$$

And our quantity of interest is $\beta$. The code for the regression models we run are as follows:

```
mod <- felm(outc ~ time+treat+DID|0|0|CensusSectors,
            data = dataframe)
```

We use the `lfe` package to estimate the models, as it facilitates the use of cluster-robust standard errors.

#### C.11 Standard Errors

We calculated robust standard errors using the `vcovHC` function in the package `sandwich`. The standard error is the same as the Stata's *comma-robust.*

#### C.12 Attrition

The attrition reports have been analysed in the data analysis section of this appendix.

#### C.13 Other Information

##### C.13.1 IRB

We received IRB from the New York University and Fundação Getulio Vargas.

- NYU IRB number: IRB-FY2017-17
- FGV IRB number: IRB-01/2017

##### C.13.2 Pre-Registration

The study was pre-registered using the EGAP pre-registry tool (#20180504AA).

##### C.13.3 Funding

We received funding from the FGV Applied Research Grant Office (RPCAP-FGV). The funding body did not interfere in the study design or implementation.

##### C.13.4 Replication Dataset

Replication materials are available at https://github.com/danilofreire/incentives-healthcare.

##### C.13.5 Acknowledgements

This research benefited from the contribution of the following research assistants:

- Guilherme Fasolin, Giovanna França, Phillipe Guedon, Natalia Liberato, Fatima Portella, Catarina Roman, Leticia Santana, Larissa Santos, and Vitor Sion.

We are very thankful for their help before, during, and after the field experiment.

##### C.13.6 Parcial Implementation and Possible Threats to the Identification

We had one substantial problem during the field, with the partial implementation of the territorial assignment in the control group. The coordinator of the control group base changed the territorial assignment because of pressure from the participants that had to work farther from the base. The coordinator assigned pair for the surroundings of the base. This problem may potentially threat the territorial comparability of the cases, but to ensure that this is not confounding our results, we used propensity-score matching to re-weight these areas. After the matching, the sectors seems comparable, and we are confident that the re-weight allows a comparable territorial assignment.

### D Session Information

```
sessionInfo()
```

```
## R version 4.0.2 (2020-06-22)
## Platform: x86_64-apple-darwin17.0 (64-bit)
## Running under: macOS Catalina 10.15.7
##
```

```
## Matrix products: default
## BLAS:   /Library/Frameworks/R.framework/Versions/4.0/Resources/lib/libRblas.dylib
## LAPACK: /Library/Frameworks/R.framework/Versions/4.0/Resources/lib/libRlapack.dylib
##
## locale:
## [1] en_US.UTF-8/en_US.UTF-8/en_US.UTF-8/C/en_US.UTF-8/en_US.UTF-8
##
## attached base packages:
## [1] parallel  stats     graphics  grDevices
## [5] utils     datasets  methods   base
##
## other attached packages:
##  [1] lfe_2.8-6          Matrix_1.3-2
##  [3] kableExtra_1.3.4  robustHD_0.6.1
##  [5] perry_0.2.0       robustbase_0.93-7
##  [7] WeightIt_0.11.0   gridExtra_2.3
##  [9] maptools_1.0-2    stargazer_5.2.2
## [11] sandwich_3.0-0    estimatr_0.30.2
## [13] lmtest_0.9-38     zoo_1.8-8
## [15] ggthemes_4.2.4    readxl_1.3.1
## [17] rgdal_1.5-23      sp_1.4-5
## [19] gpclib_1.5-6      sf_0.9-7
## [21] magrittr_2.0.1    ggmap_3.0.0
## [23] forcats_0.5.1     stringr_1.4.0
## [25] dplyr_1.0.4       purrr_0.3.4
## [27] readr_1.4.0       tidyr_1.1.2
## [29] tibble_3.0.6      ggplot2_3.3.3
## [31] tidyverse_1.3.0   rmarkdown_2.7
## [33] nvimcom_0.9-106
##
## loaded via a namespace (and not attached):
##  [1] colorspace_2.0-0
##  [2] rjson_0.2.20
##  [3] ellipsis_0.3.1
##  [4] class_7.3-18
##  [5] fs_1.5.0
##  [6] rstudioapi_0.13
```

```
##  [7] lubridate_1.7.9.2
##  [8] xml2_1.3.2
##  [9] codetools_0.2-18
## [10] splines_4.0.2
## [11] knitr_1.31
## [12] texreg_1.37.5
## [13] Formula_1.2-4
## [14] jsonlite_1.7.2
## [15] CBPS_0.21
## [16] broom_0.7.5.9000
## [17] dbplyr_2.1.0
## [18] png_0.1-7
## [19] compiler_4.0.2
## [20] httr_1.4.2
## [21] backports_1.2.1
## [22] assertthat_0.2.1
## [23] cli_2.3.0
## [24] htmltools_0.5.1.1
## [25] tools_4.0.2
## [26] gtable_0.3.0
## [27] glue_1.4.2
## [28] tinytex_0.29
## [29] Rcpp_1.0.6
## [30] cellranger_1.1.0
## [31] vctrs_0.3.6
## [32] svglite_1.2.3.2
## [33] iterators_1.0.13
## [34] xfun_0.21
## [35] ps_1.5.0
## [36] rvest_0.3.6
## [37] lifecycle_1.0.0
## [38] DEoptimR_1.0-8
## [39] MASS_7.3-53.1
## [40] scales_1.1.1
## [41] hms_1.0.0
## [42] yaml_2.2.1
## [43] gdtools_0.2.3
```

```
## [44] stringi_1.5.3
## [45] foreach_1.5.1
## [46] e1071_1.7-4
## [47] shape_1.4.5
## [48] RgoogleMaps_1.4.5.3
## [49] rlang_0.4.10
## [50] pkgconfig_2.0.3
## [51] systemfonts_1.0.1
## [52] bitops_1.0-6
## [53] evaluate_0.14
## [54] lattice_0.20-41
## [55] tidyselect_1.1.0
## [56] plyr_1.8.6
## [57] R6_2.5.0
## [58] generics_0.1.0
## [59] DBI_1.1.1
## [60] pillar_1.4.7
## [61] haven_2.3.1
## [62] foreign_0.8-81
## [63] withr_2.4.1
## [64] units_0.6-7
## [65] survival_3.2-7
## [66] nnet_7.3-15
## [67] modelr_0.1.8
## [68] crayon_1.4.1
## [69] KernSmooth_2.23-18
## [70] jpeg_0.1-8.1
## [71] grid_4.0.2
## [72] reprex_1.0.0
## [73] digest_0.6.27
## [74] classInt_0.4-3
## [75] webshot_0.5.2
## [76] xtable_1.8-4
## [77] numDeriv_2016.8-1.1
## [78] MatchIt_4.1.0
## [79] munsell_0.5.0
## [80] glmnet_4.1-1
```

```
## [81] viridisLite_0.3.0
```
